# Individualised Profiling of White Matter Organisation in Moderate-to-Severe Traumatic Brain Injury Patients Using TractLearn: A Proof-of-Concept Study

**DOI:** 10.1101/2022.03.03.22271839

**Authors:** Adam Clemente, Arnaud Attyé, Félix Renard, Fernando Calamante, Alex Burmester, Phoebe Imms, Evelyn Deutscher, Hamed Akhlaghi, Paul Beech, Peter H Wilson, Govinda Poudel, Juan F Dominguez D, Karen Caeyenberghs

**Affiliations:** Healthy Brain and Mind Research Centre, School of Behavioural, Health and Human Sciences, Faculty of Health Sciences, Australian Catholic University, Melbourne, Victoria, Australia; Cognitive Neuroscience Unit, School of Psychology, Deakin University, Geelong, Victoria, Australia; CNRS LPNC UMR 5105, University of Grenoble Alpes, Grenoble, France; School of Biomedical Engineering, The University of Sydney, Sydney, New South Wales 2006, Australia; Sydney Imaging – The University of Sydney, Sydney, Australia; Emergency Department, St. Vincent’s Hospital, University of Melbourne, Melbourne, Victoria, Australia; Department of Psychology, Faculty of Health, Deakin University; Department of Radiology and Nuclear Medicine, The Alfred Hospital, Melbourne, Victoria, Australia; Mary MacKillop Institute for Health Research, Faculty of Health Sciences, Australian Catholic University, Melbourne, Victoria, Australia

**Author notes:** Corresponding author: Dr Adam Clemente, Healthy Brain and Mind Research Centre, Australian Catholic University, Level 3, 115 Victoria Parade, Fitzroy, Melbourne VIC 3065. Shared Senior Authors (Juan F Dominguez D, Karen Caeyenberghs).

**Keywords:** traumatic brain injury, fixel-based analysis, diffusion MRI, white matter, personalised medicine

## Abstract

Approximately 65% of moderate-to-severe traumatic brain injury (m-sTBI) patients present with poor long-term behavioural outcomes, which can significantly impair activities of daily living. Numerous diffusion-weighted MRI studies have linked these poor outcomes to decreased white matter integrity of several commissural tracts, association fibres and projection fibres in the brain. However, these studies focused on group-based analyses, which are unable to deal with the substantial between-patient heterogeneity in m-sTBI. As a result, there is increasing interest in conducting individualised neuroimaging analyses. Here, we generated a detailed subject-specific characterisation of microstructural organisation of white matter tracts in 5 chronic patients with m-sTBI (29 – 49y, 2 females). We developed an imaging analysis framework using fixel-based analysis and TractLearn to determine whether the values of fibre density of white matter tracts at the individual patient level deviate from the healthy control group (*n* = 12, 8F, *M*_age_=35.7y, age range 25 – 64y). Our individualised analysis confirmed unique white matter profiles, and the heterogeneous nature of m-sTBI to properly characterise the extent of brain abnormality. Future studies incorporating clinical data, as well as utilising larger reference samples and examining the test-retest reliability of the fixel-wise metrics are warranted. This proof-of-concept study suggests that these resulting individual profiles may assist clinicians in planning personalised training programs for chronic m-sTBI patients, which is necessary to achieve optimal behavioural outcomes and improved quality of life.

---

Numerous diffusion-weighted MRI (dMRI) studies have demonstrated that moderate to severe traumatic brain injury (m-sTBI) is associated with structural white matter changes in association/projection/commissural fibre bundles, which play an important role in long-term behavioural deficits (for reviews and meta-analyses, see Roberts, Anderson, & Husain, 2014; Wallace et al., 2018). For example, Kraus et al. (2007) revealed reduced fractional anisotropy (FA) values in 13 major white matter tracts (e.g., superior longitudinal fasciculus, genu of corpus callosum) in m-sTBI patients, compared to controls. These reduced values of white matter organisation were associated with poor performance on a range of cognitive tasks (i.e., executive function, attention, and memory).

Despite these promising findings, the studies performed group-based analyses, which fail to reflect changes at an individual level. Indeed, increasing interest on group-wise comparisons on dMRI metrics (i.e., *N* patients vs *M* controls) has been questioned, and the merits of individual profiling (i.e., 1 patient vs *M* controls) extolled (Attye et al., 2021; Chamberland et al., 2021; Jolly et al., 2020; Lv et al., 2021). The goal being to enhance the characterisation of brain alterations of clinically heterogeneous groups rather than just focus on the abnormalities common to the group. Lv and colleagues (2021) investigated 45 white matter tracts in individual schizophrenia patients (*N*=322) in comparison to a group of healthy controls (*N*=195), revealing widespread but highly heterogeneous deviations in FA across all tracts across the participants. In addition, Chamberland et al. (2021) revealed large idiosyncrasies in white matter microstructure abnormalities across children with copy number variants, highlighting the need for individualised profiles. Taken together, comparison of average group differences has tended to obscure between-patient heterogeneity in the topography of the lesions, type of lesion, tissue repair and recovery mechanisms, limiting our understanding of the aetiology of TBI and identification of effective rehabilitation and treatment (Verdi et al., 2021).

To the best of our knowledge, only three studies so far have performed single-subject analyses in TBI patients (Attye et al., 2021; Jolly et al. 2020; Poudel et al., 2020). Jolly et al. (2020) conducted a subject vs reference group analysis for the identification of traumatic axonal injury in individuals with m-sTBI. They revealed substantial heterogeneity in tensor-based metrics (e.g., FA) across m-sTBI patients. Attye et al. (2021) introduced a novel framework called TractLearn, which overcomes limitations of the standard subject versus reference group studies, because it employs more specific normative values instead of computing average values of the dMRI metrics of controls as reference scores. Using TractLearn, Attye et al. (2021) detected abnormal voxels in a wide array of white matter tracts using both tensor (e.g., FA) and constrained spherical deconvolution-based (e.g., apparent fibre density, AFD) dMRI metrics in five mild TBI patients. Their findings revealed the ability of TractLearn to capture both the variability of the healthy controls (*N*=20) and the subtle quantitative alterations from a brain bundle at the voxel scale in mild TBI patients, all of which did not show visible lesions on standard MRI scans.

In the present study, our overall aim is to test the efficacy of TractLearn in m-sTBI patients with visible focal lesions. We consider TractLearn over classical voxel-to-voxel methods since this method can model better the bundle’s variability (Attye et al., 2021). Our objectives are twofold. The first aim is to develop a framework for generating subject-specific brain profiles of our patients with m-sTBI in the chronic stage of injury. To this end, we will utilise the TractLearn framework (Attye et al., 2021) to determine whether the fibre density (FD) values (derived from fixel-based analysis (FBA; Raffelt et al., 2015; Raffelt et al., 2017) of white matter tracts of an individual patient deviate from a healthy reference group (N=12). We have chosen FBA over the traditional diffusion tensor approach (as was done in the study of Jolly et al. 2020), as FBA offers greater specificity (Dhollander et al. 2021; Raffelt et al., 2012; Liang et al., 2021; Mito et al., 2018).

## Methods

### Participants

m-sTBI participants were recruited at St Vincent’s Hospital (H.A.) in Melbourne and inclusion criteria were as follows: (1) age range 18-65 years; (2) diagnosed with m-sTBI at the time of injury based on: (i) a Glasgow Coma Scale score between 3-12 (Teasdale & Jennett, 1974); (ii) loss of consciousness longer than 30 minutes; and (iii) Post traumatic Amnesia (PTA) longer than 24 hours (Rabinowitz & Levin, 2014); (3) in the chronic phase (>6 months after injury), (4) ambulant or independently mobile at the time of recruitment; (5) no known diagnosed TBI prior to current brain injury and; (6) right-handed as defined by the Edinburgh Handedness Inventory (Oldfield, 1971).

A total of six patients with m-sTBI were recruited for this study. See Table 1 for a summary of demographic, injury, and clinical characteristics for the m-sTBI participants. The patients were compared to a reference group of 12 healthy controls (HC; age range = 25 – 64 years; *M*_age_ = 35.70 ± 11.3 years; 8 females). This reference group enabled us to meaningfully assess fibre density differences between each individual TBI patient and the HC group. Healthy controls were recruited via social contacts and were included if they met the following criteria: (1) age range 18-65 years; (2) no history of neurological or psychiatric disorders; (3) right-handed as defined by the Edinburgh Handedness Inventory scale; (Oldfield, 1971; *M* = 9.92 *SD* = 0.28). We had to exclude TBI2 from the analyses, due to severe head motion in the diffusion MRI data, therefore the final number of participants for this study was five. Written informed consent was obtained from each participant in accordance with the Helsinki declaration, and ethical approval was obtained by the St Vincent’s Hospital Human Research Ethics Committee (#250/17).

**Table 1.** Summary of Demographic, Injury, and clinical Characteristics of the six m-sTBI Participants.

| Participant ID | Age Range (years)/ sex | Time since injury (years, months) | Cause | MRI scan at time of testing, Lesion location / pathology | DAI Grade<sup>2</sup> |
| --- | --- | --- | --- | --- | --- |
| TBI1 | 41-45<br>M | 21, 0 | Car accident | Small area of encephalomalacia in the (R) precentral gyrus | 0 |
| TBI2 | 46-50<br>M | 15, 6 | Motor bike accident | Large areas of encephalomalacia involving (L, R) anterior and inferior frontal lobes, (R) lateral temporal lobe and (R) parietotemporal region extending to the (R) posterior frontal lobe. Focal T1 hypointensities in the anteromedial aspect of the (L) thalamus. Volume loss and T1 hypointensity on the anterior body and genu of the corpus callosum. | 2 |
| TBI3 | 46-50<br>F | 3, 8 | Horse riding accident | Bilateral anterior and inferior frontal encephalomalacia, (R) greater than (L), and (R) anterior temporal encephalomalacia. Small deep white matter T2 hyperintensities medial (R) parietal lobe. Small focal T1 hypointensity in the anterior body of the corpus callosum. | 2 |
| TBI4 | 26-30<br>F | 15, 6 | Horse riding accident | Bilateral inferior frontal and (L) anterior temporal encephalomalacia. Small area of encephalomalacia (L) superior frontal gyrus. (R) frontal burr hole with underlying ventricular drain tract. | 1 |
| TBI5 | 46-50<br>M | 18, 1 | Car accident | Two small nonspecific deep white matter T2 hyperintensities in the (R) parietal lobe (within normal limits for age). | 0 |
| TBI6 | 26-30<br>F | 5, 10 | Horse riding accident | Small T1 hypointensity in splenium of corpus callosum. Scattered punctate T2 hyperintensities in both cerebral hemispheres (approx. 6). | 2 |

<sup>1</sup> Grading of Diffuse Axonal Injury (DAI) in accordance with Adams et al. (1989) grading: a "0" grade indicates no confirmed DAI present; "1" there is histological evidence of axonal injury in the white matter of the cerebral hemispheres, the corpus callosum, the brainstem and, less commonly, the cerebellum; "2" there is also a focal lesion in the corpus callosum; "3" there is additionally a focal lesion in the dorsolateral quadrant or quadrants of the rostral brainstem. Anatomical T1-weighted image and fluid-attenuated inversion recovery MRI scans were inspected and classified by an experienced neuroradiologist (P.B.) providing the descriptions of the lesions and DAI grading. Abbreviations: M = male; F = female;; L = left; R = right

### MRI Acquisition

Structural MRI scans were acquired on a 3T Siemens PRISMA scanner with a 64-channel head coil at Murdoch Children’s Research Institute in Melbourne, Australia. Single shell diffusion-weighted imaging was acquired using single-shot echo planar with twice-reinforced spin echo and a multi-band acceleration factor of 2 which was obtained with 70 contiguous transverse slices (FOV = 260 × 260 mm^2^, voxel size = 2.3mm isotropic, TR = 3500ms, TE = 67ms, A>>P, and TA=6 minutes 17 seconds. A high angular resolution diffusion imaging (HARDI; Frank, 2001) gradient scheme was applied in 64 non-collinear gradient directions, maximum b-value of 3000s/mm^2^, and seven interleaved b0 images. A pair of anterior–posterior (AP) and posterior–anterior (PA) reverse phase-encoded images were also collected to correct for geometric distortions (TA = 50 seconds each).

In addition, anatomical MRI scans were acquired using a T1 weighted imaging (ADNI protocol with 104 contiguous sagittal slices (A>>P, FOV = 220 × 220 mm^2^, voxel size = 1.0×1.00×1.50mm^3^, TR = 2250ms, TE = 3.07ms, flip angle = 9º, and TA = 5.48min) and fluid-attenuated inversion recovery (FLAIR: 176 slices; FOV = 256mm; voxel size = 0.5×0.5×0.9mm^3^; TR = 6000ms; TE = 437ms; TI = 2100ms; TA = 7:20min) scan sequence. These MRI scans were inspected and classified by an experienced neuroradiologist (P.B.) providing an overall description of the pathology as well as a DAI grade using the Adams et al. (1989) criteria (as can be seen in Table 1).

### MRI data processing

The current study uses a state-of-the-art processing pipeline (see Supplementary Material 1.0 for detailed description), which inherently deals with large focal lesions (Dhollander et al., 2021) in m-sTBI patients. See Figure 1 for a schematic overview of the pipeline.

**Figure 1.**
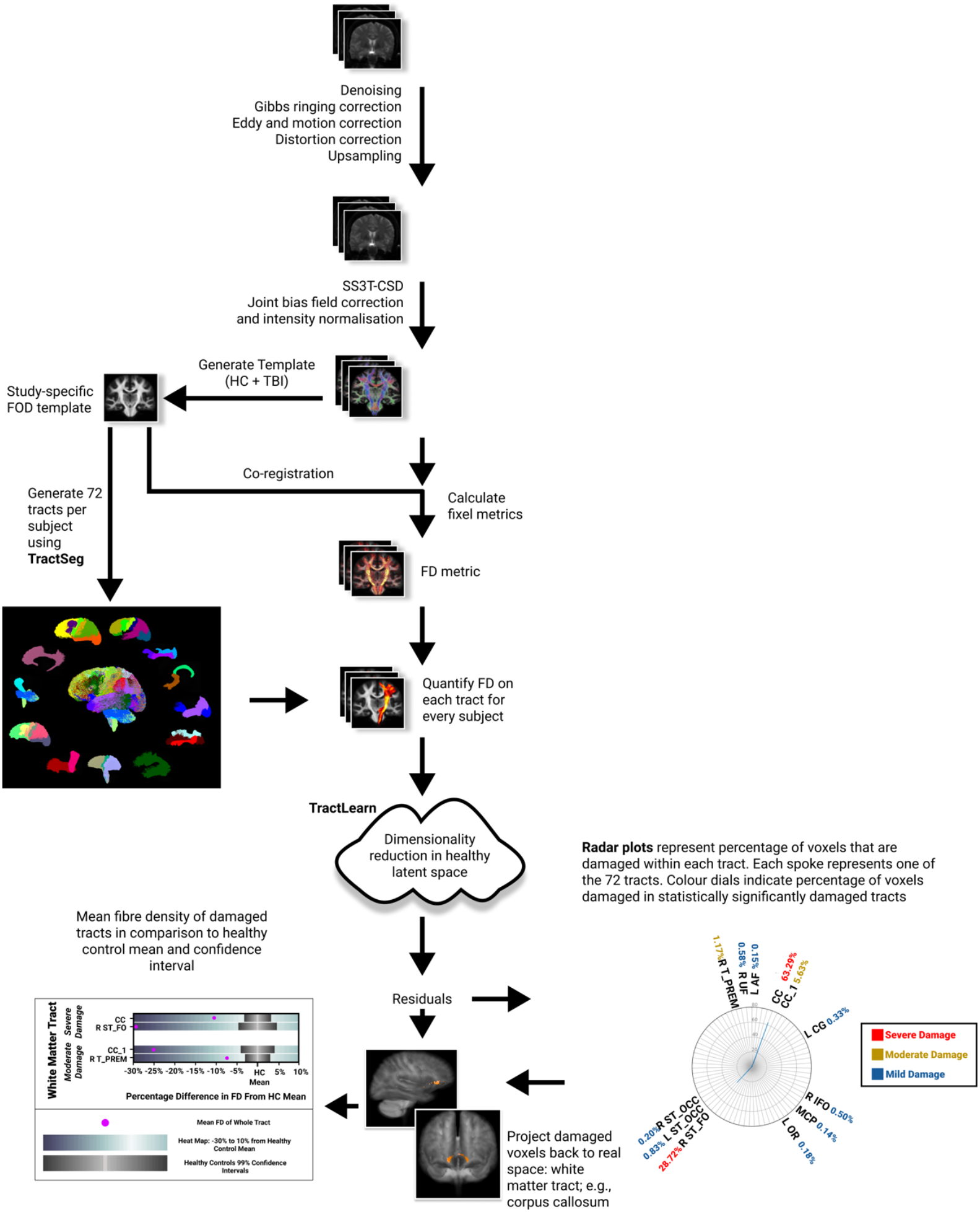
Schematic overview of MRI framework (A) Raw diffusion images are pre-processed to remove artefacts (noise, Gibbs-ringing, motion and distortion correction). (B) Single shell 3 tissue constrained spherical deconvolution is then performed on the pre-processed images followed by joint bias field correction and intensity normalisation. (C) A study-specific population template is then constructed using HC (cross-sectional data) and m-sTBI patient data. (D) For each HC and m-sTBI participant, 72 tracts are generated using TractSeg and fixel-wise fibre density (FD) is computed for each individual tract. (E) Using a manifold learning approach, TractLearn then detects abnormal voxels in fibre density for the m-sTBI patients when compared to the healthy controls, radar plots demonstrating the percentage of damaged voxels and back-projections of damaged voxels in population template space are finally computed. (F) Heat maps of the mean FD of damaged tracts in relation to the mean FD of each tract of the healthy control group.

### Individualised profiling

To develop individualised profiles, our analysis was two-fold. In the first analysis, we used the TractLearn toolbox (Attye et al., 2021; https://github.com/GeodAIsics/TractLearn-WholeBrain). The TractLearn toolbox was utilised to identify damaged white matter tracts from the 72 tracts delineated via TractSeg (Wasserthal et al., 2018; 2019). TractLearn detects abnormal voxels for each tract in each patient, by using a manifold learning approach to determine which voxels deviate from normative healthy controls (Attye et al., 2021). In particular, we examined the damage of the fixel-wise FD metric. We computed the percentage of damaged voxels within each tract delineated via TractSeg, after stringent Bonferroni corrections. This analysis provides us with an estimate of the extent of damage in FD at the individual subject level. We categorise the severity of the extent of damage of FD for each tract as either “severe” (>10% damaged voxels), “moderate” (1-10% damaged voxels), or “mild” (<1% damaged voxels). For visualisation, these damaged voxels were then projected back to population template space. (See Supplementary Figure 1 for an example radar plot and the full list of abbreviations of tracts.)

In the second analysis, we calculated the mean FD of the whole tract for each that were moderate and severely damaged detected through TractLearn for each individual patient. FD was also calculated for the reference HC group to generate reference points for comparisons against each individual TBI patient, in which 99% confidence intervals (CIs) and the mean FD of each tract of the HCs were calculated. Finally, we classified the profiles of the patients in line with previous normative work by Lv et al. (2021), whereby FD of the tract of the patient is classified as either “normal” (if the FD value of the tract falls within the 99% CI of the HC), “supra-normal” (greater than the 99% CI of the HC), or “infra-normal” (lower than the 99% CI of the HC). This provides an overall measure representative of FD loss in tracts relative to controls.

## Results

TBI1 presented with the highest number of injured tracts (39 tracts including 3 severe, 18 moderate, 18 mild damage), most notably in the corpus callosum (segments 1, 4, 5 - moderate), thalamo postcentral (severe), fornix (severe), and CST (moderate), with some of the tracts including at least 10% damaged voxels. The calculated percentage from the HC FD means for these tracts ranged between −10 to −35%, which were classified as infra-normal. TractLearn results for TBI3 showed a significant number of damaged voxels in the corpus callosum (whole CC, segment 1; severe and moderate damage), left cingulum, left arcuate fasciculus (both mild). The moderate and severely damaged tracts, include the corpus callosum (whole CC, segment 1), right thalamo-premotor, and right striato-fronto-orbital tract which revealed an 8-30% FD loss (infra-normal) relative to the reference group. The radar plot of TractLearn results for TBI4 showed a higher number of damaged voxels (in 8 tracts) presenting with significantly altered FD values in the corticospinal tracts (mild), fornix (severe), right anterior thalamic radiation (mild), corpus callosum (whole CC, moderate), left fronto-pontine tract (mild), and left striato-occipital tract (mild). The damage in the corpus callosum and fornix showed an infra-normal 10-20% FD decrease relative to controls. The TractLearn results for TBI5 showed especially a high number of damaged voxels presenting with significant altered FD in 20 white matter tracts, including the corpus callosum (whole CC moderate damage, mild damage in segments 5-7), optic radiations (mild to moderate damage), striato-occipital bundles (mild-moderate damage), corticospinal tract (moderate) and thalamic post central bundles (moderate). Apart from the right thalamic postcentral bundle, the moderate-and-severely damaged tracts could be classified as infra-normal (5-30% FD loss) compared to the CI of the HCs. TBI6 patient presented only with damaged voxels in the left Striato-occipital fibre bundle (mild). Example TractLearn radar plots are presented in Figure 2, and FD heatmaps are presented in Figure 3. In addition, the back projections of moderate to severely damaged tracts allowed to identify the location of altered voxels within each bundle using the FD, as illustrated in Figure 4 for TBI1 and TBI3 patients.

**Figure 2.**
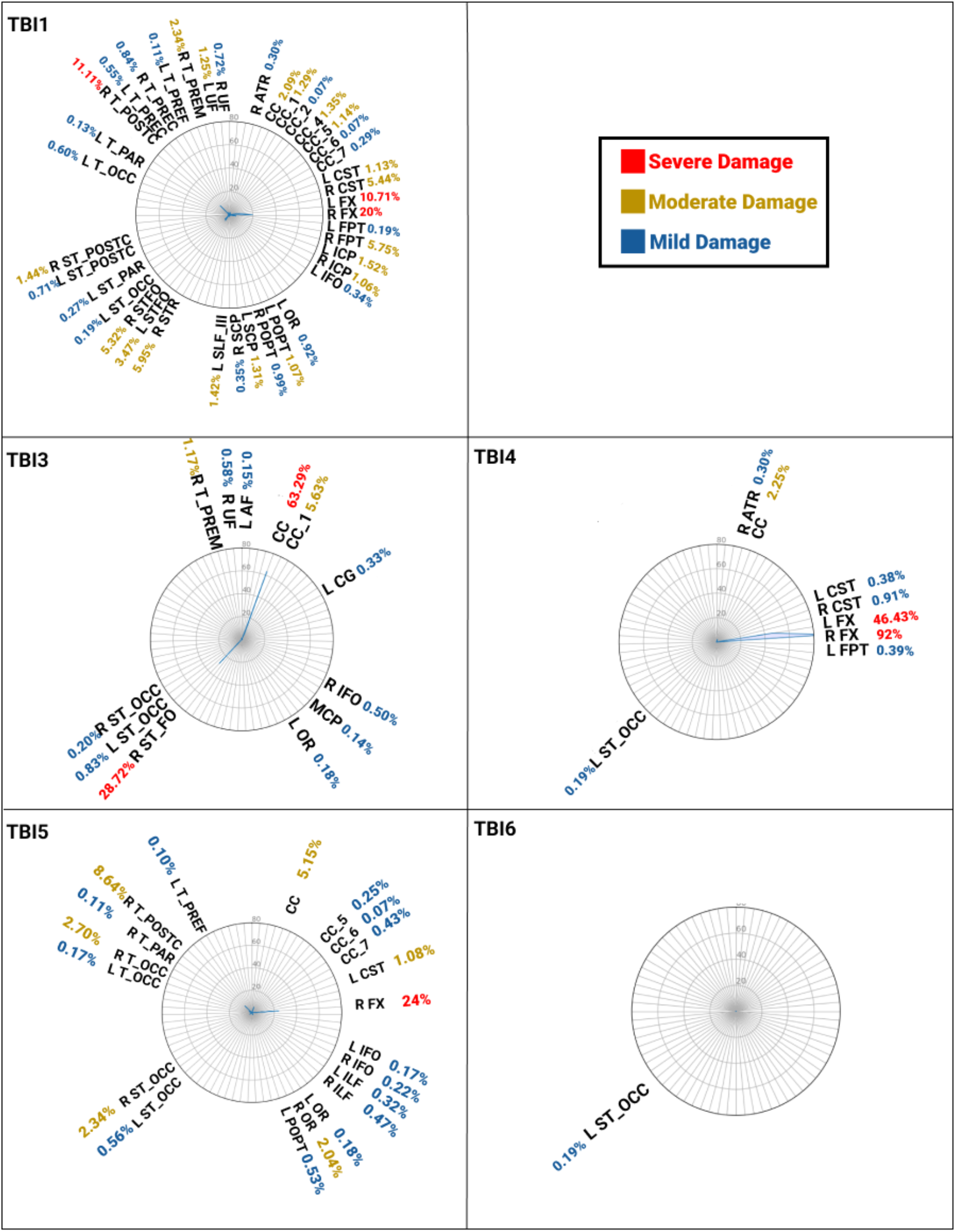
Individualised profiles of all TBI patients, presenting the percentage of damaged voxels of FD in various tracts using TractLearn. Numbers of damage denote the severity of the damage; blue = mild, yellow = moderate, red = severe.

**Figure 3.**
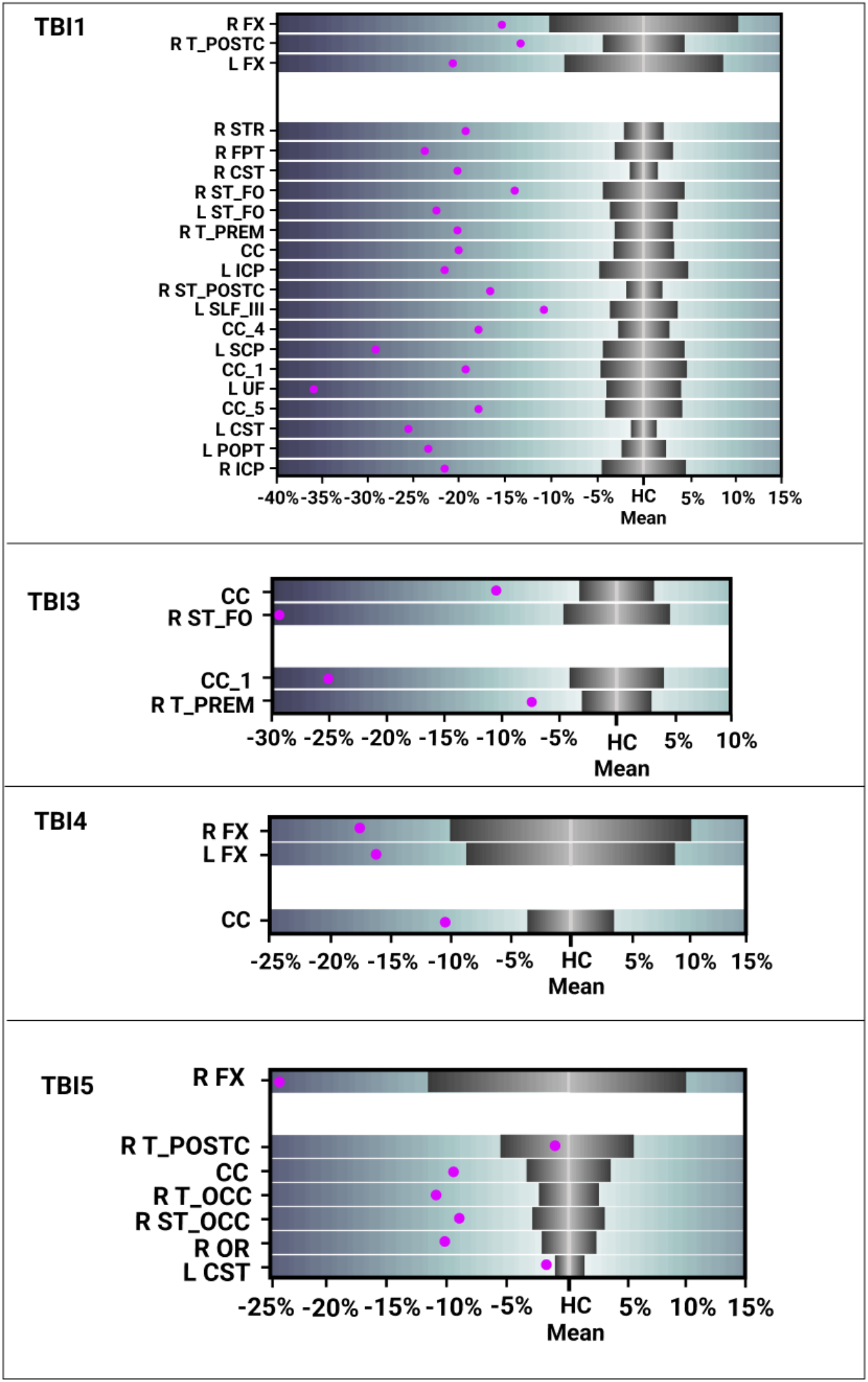
Individualised profiles of m-sTBI patients present FD heatmaps. Each heatmap shows the mean FD of the severe (top panel) and moderately (bottom panel) damaged tracts. Tracts are listed from the highest number of damaged voxels to the lowest. For each heatmap, the X-axis represents the percentage difference in FD from the healthy control mean. Grey heat maps in the centre represent the 99% healthy control confidence intervals.

**Figure 4.**
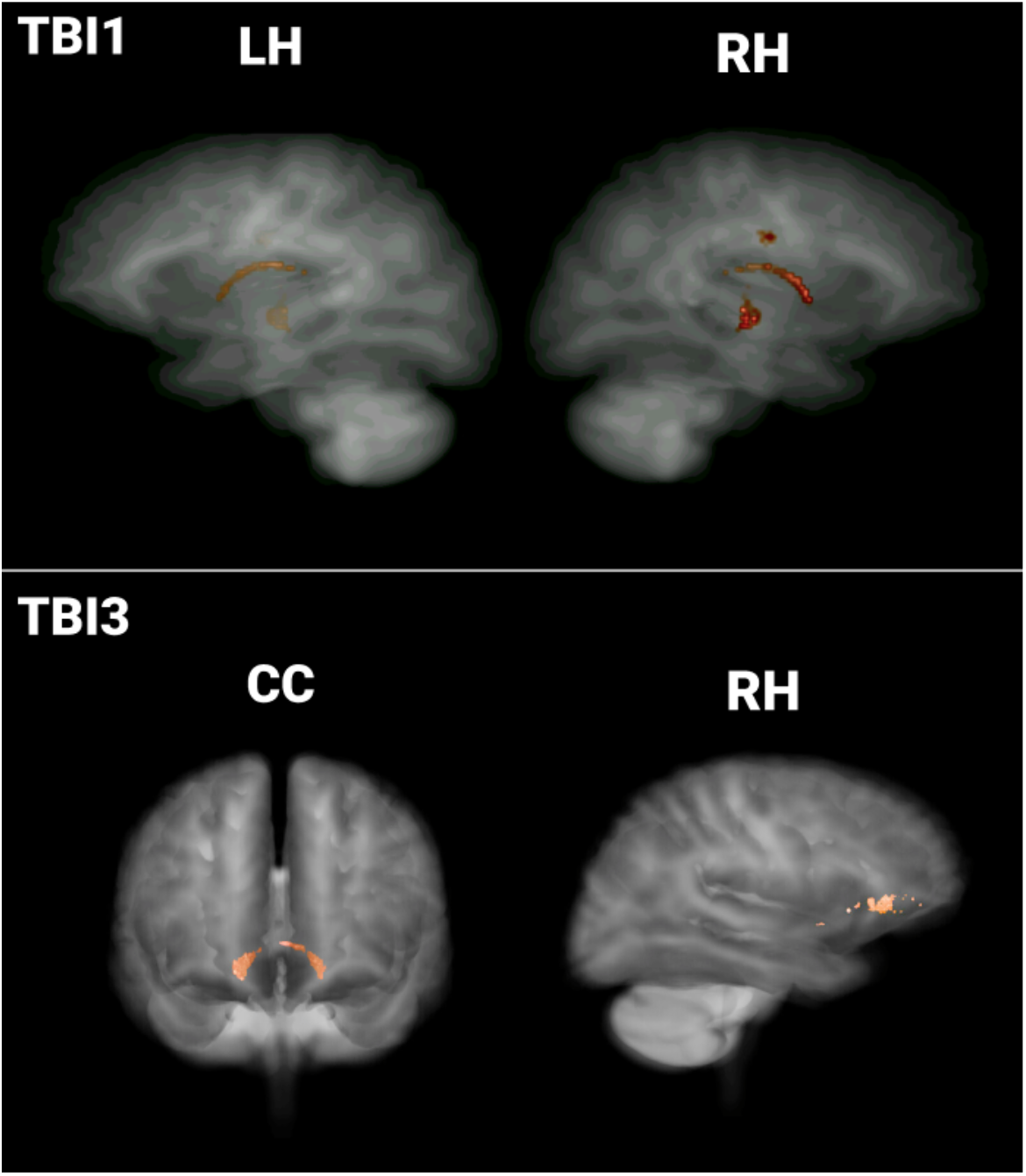
Individualised profile of TBI1 and TBI3, presenting the back-projection visualisations of the damaged voxels of the severely damaged tracts projected back into population template space. LH = left hemisphere; RH = right hemisphere, CC = corpus callosum.

## Discussion

The present study provides a proof-of-concept study of TractLearn in five patients with m-sTBI to detect patient-specific FD loss patterns in comparison to healthy controls. These results provided further evidence of the benefits of characterising abnormalities at the patient level when dealing with highly heterogeneous populations. First, we will discuss our observations from these individualised profiles followed by discussion of key ideas to improve individualised profiles of tract-specific white matter.

### Individualised profiling of white matter microstructure

These individual profiles showed a wide variation across participants from one damaged tract (i.e., TBI6) to 39 damaged tracts (i.e., TBI1). Interestingly, the secondary analysis of FD loss relative to controls showed that the majority of tracts were damaged, some up to 40% difference in FD relative to controls. This mean FD plots provides further insights into the large heterogeneous damage across m-sTBI patients. This reflects that white matter organisation in m-sTBI is sensitive to damage relative to controls in high rates, which may be a biomarker of behavioural deficits. These results highlight the variability and heterogeneity that is inherent in m-sTBI patients, which is consistent with reports from previous group-based studies (Han et al., 2017; Strangman et al., 2010; Verhelst et al., 2019). Interestingly, there were tracts which seem to be vulnerable and were present across multiple participants, this includes damage to various segments of the corpus callosum, fornix, and thalamo post central tracts, which is in line with previous research (Caeyenberghs et al., 2011; Wallace et al., 2018). For example, these findings are in line with previous dMRI studies utilising tensor-based metrics, showing the CC as a vulnerable tract to long-term damage following m-sTBI (Caeyenberghs et al., 2011). This is common in m-sTBI patients in the chronic phase, who are prone to neurodegeneration of the white matter (Poudel et al., 2020). Mechanisms of these white matter damage observed is most likely the result of secondary injury mechanisms, such as (Wallerian) degeneration (Koliatsos & Alexandris, 2019). Further studies are required to correlate the severity of white matter damage with patients’ neurocognitive profiles.

### State-of-the art processing pipeline and implications of single-subject profiling

Our patients showed large focal lesions, representative of m-sTBI. However, using single-shell 3 tissue constrained spherical deconvolution (SS3T-CSD; Dhollander and Connelly, 2016; Dhollander, Mito, Raffelt, & Connelly, 2019) we were able to reconstruct the majority of the white matter tracts in a robust manner. This approach utilises group-average response functions for white matter, grey matter, and cerebrospinal fluid tissue types (Dhollander et al., 2016; Dhollander, Raffelt, & Connelly, 2016). These response functions are tissue-specific signal models that allow tissue signal fraction estimation of grey matter and cerebrospinal fluid signals as well as white matter fibre orientation distributions (FODs) through spherical deconvolution analysis. This allows for clear delineation of white matter fibre-bundles for TractLearn analysis. Computing white matter FODs using SS3T-CSD has previously been suggested to be able to inherently deal with lesions (Dhollander et al., 2020; Egorova et al., 2020; Gajamange et al. 2018; Gottlieb et al., 2020). In addition, we also performed whole brain analysis at baseline through the use of TractSeg utilising the fibre-specific FD metric of the FBA framework, which overcome drawbacks of tensor-based metrics (i.e., do not account for crossing fibres) used in previous studies such as Jolly et al. (2020). Of note, computing FD could not be done in TBI2 due to excessive motion during the DWI sequence. In order for advanced analyses, control for subject motion is essential, an issue that can be of particular importance in TBI patients, whom can be less cooperative during MRI scanning. This could be achieved through preparation in a dummy scanner, more online tracking of motion during scanning, and re-running the protocol if excessive motion is detected.

Our single-subject profiles provide a subject-specific categorisation of damaged white matter tracts in m-sTBI patients in the chronic phase of their injury. TractLearn was used to detect the damaged tracts, which relies on a manifold learning approach (Attye et al., 2021) This framework (as opposed to deep-learning method) for single-subject anomaly detection has two advantages over classic statistics methods:

1/Decreasing the number of false positive findings by capturing the anatomical variability and contrast variation from the healthy atlas. Although a benefit of manifold learning (compared with deep learning) is that it does not rely on the need of large number of subjects, future studies could benefit from using slightly larger control groups for reference groups (*N*>50).

2/Decreasing the number of false negatives by providing a nonlinear global analysis of all voxels in brain bundles, as opposed to a classic statistical method where all voxels are analysed independently.

We suggest that this very advanced method of lesion detection has accurately identified extensive fibre bundles damage in TBI patients in comparison with classic methods. However, in the absence of gold-standard protocols (i.e., methods that are widely accepted for clinical use), it is difficult to compare models. Overall, with further extensions (e.g., incorporating behavioural data) and validations in future studies, our profiles may assist clinicians in implementing personalised training programs in m-sTBI patients. For example, TBI4 with the damaged voxels in sensorimotor tracts can benefit from a motor training program (such as a balance training program) to improve motor functioning and restore white matter organisation (Liang et al., 2021).

### Limitations and future studies

There are some limitations in this study that should be considered for future research. Due to small sample size of the reference group, this study should be considered as a proof-of-concept study and future studies with larger sample sizes are needed (Jolly et al., 2020; Lv et al., 2021). This larger control group is important to ensure that the secondary analysis involving mean FD relative to controls is representative of the true population and indeed maps the heterogeneity in age, sex, and other sample characteristics. Next, although Attye et al. (2021) demonstrated high test-retest reliability for TractSeg and TractLearn, future studies should examine test-retest reliability across different scanners (vendor, software, strength) and data acquisition (Grech-Sollars et al., 2015; Karayumak et al., 2019; Mirzaalian et al., 2016; Schmeel, 2019; Tax et al., 2019). Also, the development of a longitudinal pipeline to track the progression of damaged tracts over time or after a specific training program may have further implications for personalised treatment regimes and has not yet been attempted. Conducting TractLearn in a longitudinal analysis is possible and has the potential to overcome these drawbacks. The next key important step is to incorporate behavioural data to further understand the biomarker implication of structural changes in the white matter in TBI patients. Linking white matter profiles to clinical behavioural profiles which have been shown to correlate with damaged tracts may also provide added value for personalised training and is therefore an important step to translate the basic science into the clinical practice. This may eventually lead to tailored training with details provided at baseline or at the end of a training protocol, leading to the potential for greater health options for heterogeneous populations.

## Conclusion

Overall, this study presents a novel *individualised profiling* framework, extending the use of TractLearn in m-sTBI patients, with the extension of further profiles of the damage tracts relative to controls. This approach captures the heterogeneity within individual m-sTBI patients and is therefore beneficial in clinically heterogeneous populations. This study extends on the recent papers on single-subject profiling (e.g., Attye et al., 2021; Chamberland et al., 2021; Jolly et al., 2020; Lv et al., 2021) by developing further insight into damaged tracts, as well as using specific FBA-framework based FD measures. With further validation of this approach, single-subject profiles may eventually assist clinicians with more quantitative information about the changes in the brain structure, potentially augmenting diagnostic and/or treatment decisions made with respect to individual patients with m-sTBI.

## Data Availability

All data produced in the present study are available upon reasonable request to the authors

## Notes

### Competing Interest Statement

The authors have declared no competing interest.

### Funding Statement

This work was facilitated by an Australian
Catholic University Research Fund
[#902915]. Karen Caeyenberghs is supported by a
National Health and Medical Research
Council Career Development Fellowship
(APP1143816).

### Author Declarations

St Vincent's Hospital Human Research Ethics Committee of St Vincent's Hospital Melbourne gave ethical approval for this work (HREC #250/17)

## References

Adams, J. H., Doyle, D., Ford, I., Gennarelli, T. A., Graham, D. I., & McLellan, D. R. (1989). Diffuse axonal injury in head injury: Definition, diagnosis and grading. Histopathology, 15(1), 49–59. doi: 10.1111/j.1365-2559.1989.tb03040.x

Attye, A., Renard, F., Baciu, M., Roger, E., Lamalle, L., Dehail, P., … Calamante, F. (2021). TractLearn: A geodesic learning framework for quantitative analysis of brain bundles. Neuroimage, 233, 117927 doi: 10.1101/2020.05.27.20113027

Caeyenberghs, K., Leemans, A., Coxon, J., Leunissen, I., Drijkoningen, D., Geurts, M., … & Swinnen, S. P. (2011). Bimanual coordination and corpus callosum microstructure in young adults with traumatic brain injury: A diffusion tensor imaging study. Journal of Neurotrauma, 28(6), 897–913. doi: 10.1089/neu.2010.1721

Chamberland, M., Genc, S., Tax, C. M., Shastin, D., Koller, K., Raven, E. P., … & Jones, D. K. (2021). Detecting microstructural deviations in individuals with deep diffusion MRI tractometry. Nature Computational Science, 1(9), 598–606. doi: 10.1038/s43588-021-00126-8

Dhollander, T., & Connelly, A. (2016). A novel iterative approach to reap the benefits of multi-tissue CSD from just single-shell (+b=0) diffusion MRI data. In: Proceedings of the International Society for Magnetic Resonance in Medicine, 3010.

Dhollander, T., Clemente, A., Singh, M., Boonstra, F., Civier, O., Dominguez D J., … & Caeyenberghs, K. (2021). Fixel-based analysis of diffusion MRI: methods, applications, challenges and opportunities. Neuroimage, 241, 118417. doi: 10.1016/j.neuroimage.2021.118417

Dhollander, T., Mito, R., Raffelt, D., & Connelly, A. (2019). Improved white matter response function estimation for 3-tissue constrained spherical deconvolution. In: Proceedings of the International Society for Magnetic Resonance in Medicine, 555.

Dhollander, T., Raffelt, D., & Connelly, A. (2016). Unsupervised 3-tissue response function estimation from single-shell or multi-shell diffusion MR data without a co-registered T1 image. ISMRM Workshop on Breaking the Barriers of Diffusion MRI, 5.

Egorova, N., Dhollander, T., Khlif, M. S., Khan, W., Werden, E., & Brodtmann, A. (2020). Pervasive white matter fiber degeneration in ischemic stroke. Stroke, 51(5), 1507–1513. doi: 10.1161/STROKEAHA.119.028143

Frank, L. R. (2001). Anisotropy in high angular resolution diffusion-weighted MRI. Magnetic Resonance in Medicine, 45(6), 935–939. doi: 10.1002/mrm.1125

Gajamange, S., Raffelt, D., Dhollander, T., Lui, E., van der Walt, A., Kilpatrick, T., … & Kolbe, S. (2018). Fibre-specific white matter changes in multiple sclerosis patients with optic neuritis. NeuroImage: Clinical, 17, 60–68. doi: 10.1016/j.nicl.2017.09.027

Gottlieb, E., Egorova, N., Khlif, M. S., Khan, W., Werden, E., Pase, M. P., … Brodtmann, A. (2020). Regional neurodegeneration correlates with sleep-wake dysfunction after stroke. Sleep, 43(9), zsaa054. doi: 10.1093/sleep/zsaa054

Grech-Sollars, M., Hales, P. W., Miyazaki, K., Raschke, F., Rodriguez, D., Wilson, M., … Gwilliam, M. N. (2015). Multi-centre reproducibility of diffusion MRI parameters for clinical sequences in the brain. NMR in Biomedicine, 28(4), 468–485. doi: 10.1002/nbm.3269

Han, K., Davis, R. A., Chapman, S. B., & Krawczyk, D. C. (2017). Strategy-based reasoning training modulates cortical thickness and resting-state functional connectivity in adults with chronic traumatic brain injury. Brain and Behaviour, 7(5), 1–27. doi:10.1002/brb3.687

Jolly, A. E., Bălăeţ, M., Azor, A., Friedland, D., Sandrone, S., Graham, N. S., … Sharp, D. J. (2020). Detecting axonal injury in individual patients after traumatic brain injury. Brain, awaa372. doi: 10.1093/brain/awaa372

Karayumak, S. C., Bouix, S., Ning, L., James, A., Crow, T., Shenton, M., … Rathi, Y. (2019). Retrospective harmonization of multi-site diffusion MRI data acquired with different acquisition parameters. Neuroimage, 184, 180–200. doi: 10.1016/j.neuroimage.2018.08.073

Koliatsos, V. E., & Alexandris, A. S. (2019). Wallerian degeneration as a therapeutic target in traumatic brain injury. Current Opinion in Neurology, 32(6), 786. doi: 10.1097/WCO.0000000000000763

Kraus, M. F., Susmaras, T., Caughlin, B. P., Walker, C. J., Sweeney, J. A., & Little, D. M. (2007). White matter integrity and cognition in chronic traumatic brain injury: a diffusion tensor imaging study. Brain, 130(10), 2508–2519. doi: 10.1093/brain/awm216

Liang, X., Yeh, C. H., Poudel, G., Swinnen, S. P., & Caeyenberghs, K. (2021). Longitudinal fixel-based analysis reveals restoration of white matter alterations following balance training in young brain-injured patients. NeuroImage: Clinical, 30, 102621. doi: 10.1016/j.nicl.2021.102621

Lv J., Di Biase, M., Cash, R. F., Cocchi, L., Cropley, V. L., Klauser, P., … & Zalesky, A. (2021). Individual deviations from normative models of brain structure in a large cross-sectional schizophrenia cohort. Molecular Psychiatry, 26(7), 3512–3523. doi: 10.1038/s41380-020-00882-5

Mirzaalian, H., Ning, L., Savadjiev, P., Pasternak, O., Bouix, S., Michailovich, O., … George, M. S. (2016). Inter-site and inter-scanner diffusion MRI data harmonization. NeuroImage, 135, 311–323. doi: 10.1016/j.neuroimage.2016.04.041

Mito, R., Raffelt, D., Dhollander, T., Vaughan, D. N., Tournier, J. D., Salvado, O., … Connelly, A. (2018). Fibre-specific white matter reductions in Alzheimer’s disease and mild cognitive impairment. Brain, 141(3), 888–902. doi: 10.1093/brain/awx355

Oldfield, R. C. (1971). The assessment and analysis of handedness: The Edinburgh inventory. Neuropsychologia, 9(1), 97–113. doi: 10.1016/0028-3932(71)90067-4

Poudel, G. R., Dominguez D J. F., Verhelst, H., Vander Linden, C., Deblaere, K., Jones, D. K., … & Caeyenberghs, K. (2020). Network diffusion modeling predicts neurodegeneration in traumatic brain injury. Annals of Clinical and Translational Neurology, 7(3), 270–279. doi: 10.1002/acn3.50984

Rabinowitz, A., & Levin, H. (2014). Cognitive sequelae of traumatic brain injury. Psychiatric Clinics of North America, 37(1), 1–11. doi: 10.106/j.psc.2013.11.004

Raffelt, D. A., Smith, R. E., Ridgway, G. R., Tournier, J. D., Vaughan, D. N., Rose, S., … Connelly, A. (2015). Connectivity-based fixel enhancement: Whole-brain statistical analysis of diffusion MRI measures in the presence of crossing fibres. Neuroimage, 117(1), 40–55. doi: 10.1016/j.neuroimage.2015.05.039

Raffelt, D. A., Tournier, J. D., Smith, R. E., Vaughan, D. N., Jackson, G., Ridgway, G. R., Connelly, A. (2017). Investigating white matter fibre density and morphology using fixel-based analysis. Neuroimage, 144(A), 58–73. doi: 10.1016/j.neuroimage.2016.09.029

Raffelt, D., Tournier, J. D., Crozier, S., Connelly, A., & Salvado, O. (2012). Reorientation of fibre orientation distributions using apodized point spread functions. Magnetic Resonance in Medicine, 67(3), 844–855. doi: 10.1002/mrm.23058

Roberts, R. E., Anderson, E. J., & Husain, M. (2013). White matter microstructure and cognitive function. The Neuroscientist, 19(1), 8–15. doi: 10.1177/1073858411421218

Schmeel, F. C. (2019). Variability in quantitative diffusion-weighted MR imaging (DWI) across different scanners and imaging sites: Is there a potential consensus that can help reducing the limits of expected bias?. European Radiology, 29, 2243–2245. doi: 10.1007/s00330-018-5866-4

Strangman, G. E., O’Neil-Pirozzi, T. M., Supelana, C., Goldstein, R., Katz, D. I., & Glenn, M. B. (2010). Regional brain morphometry predicts memory rehabilitation outcome after traumatic brain injury. Frontiers in Human Neuroscience, 4, 182. doi: 10.3389/fnhum.2010.00182

Tax, C. M., Grussu, F., Kaden, E., Ning, L., Rudrapatna, U., Evans, C. J., … Ghosh, A. (2019). Cross-scanner and cross-protocol diffusion MRI data harmonisation: A benchmark database and evaluation of algorithms. Neuroimage, 195, 285–299. doi: 10.1016/j.neuroimage.2019.01.077

Teasdale, G., & Jennett, B. (1974). Assessment of coma and impaired consciousness: A practical scale. The Lancet, 304(7872), 81–84. doi: 10.1016/S0140-6736(74)91639-0

Verdi, S., Marquand, A. F., Schott, J. M., & Cole, J. H. (2021). Beyond the average patient: how neuroimaging models can address heterogeneity in dementia. Brain, 144(10), 2946–2953. doi: 10.1093/brain/awab165

Verhelst, H., Giraldo, D., Vander Linden, C., Vingerhoets, G., Jeurissen, B., & Caeyenberghs, K. (2019). Cognitive training in young patients with traumatic brain injury: A fixel-based analysis. Neurorehabilitation and Neural Repair, 33(10), 813–824. doi: 10.1177/1545968319868720

Wallace, E. J., Mathias, J. L., & Ward, L. (2018). The relationship between diffusion tensor imaging findings and cognitive outcomes following adult traumatic brain injury: A meta-analysis. Neuroscience & Biobehavioral Reviews, 92, 93–103. doi: 10.1016/j.neubiorev.2018.05.023

