## Supplementary for "Individualised Profiling of White Matter Organisation in Moderate-to-Severe Traumatic Brain Injury Patients Using TractLearn: A Proof-of-Concept Study"

### Supplementary Material

#### 1.0 MRI Pre-Processing

***Pre-processing.*** MRI data was processed using MRtrix3Tissue (https://3Tissue.github.io), a fork of MRtrix3 (Tournier et al. 2019). The pre-processing steps included: denoising (Veraart et al., 2016), removal of Gibbs ringing (Kellner et al., 2016), and eddy current, motion, and susceptibility induced distortion correction with outlier replacement (Andersson & Sotiropoulous, 2016; Andersson et al., 2016; Smith et al., 2004). The pre-processed diffusion weighted images were then upsampled from 2.3 mm to 1.33 mm isotropic voxels before the computation of the upsampled brain masks.

***Fibre orientation distribution calculation.*** As our m-sTBI patients had large focal lesions, a robust pipeline was needed to control for this. We estimated the fibre orientation distributions (FOD) estimations using single-shell 3-tissue constrained spherical deconvolution (SS3T-CSD; Dhollander and Connelly, 2016). Research on patients with lesions in both MS (e.g., Gajamange et al. 2018) and stroke (e.g., Egorova et al., 2020; Gottlieb et al., 2020) have computed white matter FODs using SS3T-CSD which is suggested to be the best current method for dealing with lesions (Dhollander, Mito, Raffelt, & Connelly, 2019; Dhollander, Raffelt, & Connelly, 2016; Dhollander et al., 2021). Following this, we performed joint bias field correction and global intensity normalization (Raffelt et al., 2015).

***Fibre orientation distribution template construction.*** We generated a study specific unbiased FOD template using the FOD templates from the (1) six m-sTBI participants and; (2) the 12 cross-sectional HC participants, using linear and non-linear registration of the FOD image (Raffelt et al., 2011; Raffelt et al., 2012). This method for template construction is used in other studies profiling single-subjects (Attye et al., 2021) and is recommended on the MRtrix3 documentation (www.mrtrix3.org). This was followed by the registration of each FOD image to template space (Raffelt et al, 2012). FOD segmentation was then performed to compute fixels at both template-level and individual-level (Raffelt et al., 2015; Raffelt et al., 2017) Finally, fixels at individual-level were all reoriented to the corresponding fixels of the FOD template in order to conduct group comparisons of fixel-wise metrics (Raffelt et al., 2011).

***Fibre density calculation.*** Based on the 2 million tractograms, the FD fixel-metric was calculated in template space for each individual participant as we are interested in microstructural changes of the white matter (Raffelt et al., 2015). For the TractLearn tool, FD was converted to voxels in order for the toolbox to run as it requires a specific form of quantitative data. These statistics were utilised for further analyses.

***Tracts of interest construction.*** We used the automated TractSeg tool to delineate 72 tracts (Wasserthal, Neher, & Maier-Hein, 2018; Wasserthal, Neher, Hirjak, & Maier-Hein, 2019). This approach provides a good balance between manual delineation and automated atlas-based tracking approaches (Genc et al., 2020; Wasserthal et al., 2018; Wasserthal et al., 2019). We run this in individual space to check that tracts were not being delineated into regions of lesioned tissue. Following this, similar to recent applications of this toolbox using FBA, for each individual participant, we warped the tracts from subject space into a common population template space to be able to compare metrics along each tract more robustly (Attye et al., 2021; Genc et al., 2020). All tracts were generated using the default TractSeg pipeline for each individual participant at each time point using their individual white matter FODs which were then warped into population template space (https://github.com/MIC-DKFZ/TractSeg), apart from upgrading each tract from the default 2000 to 10,000 streamlines. These tracts were used for further analyses.

#### Supplementary Figures

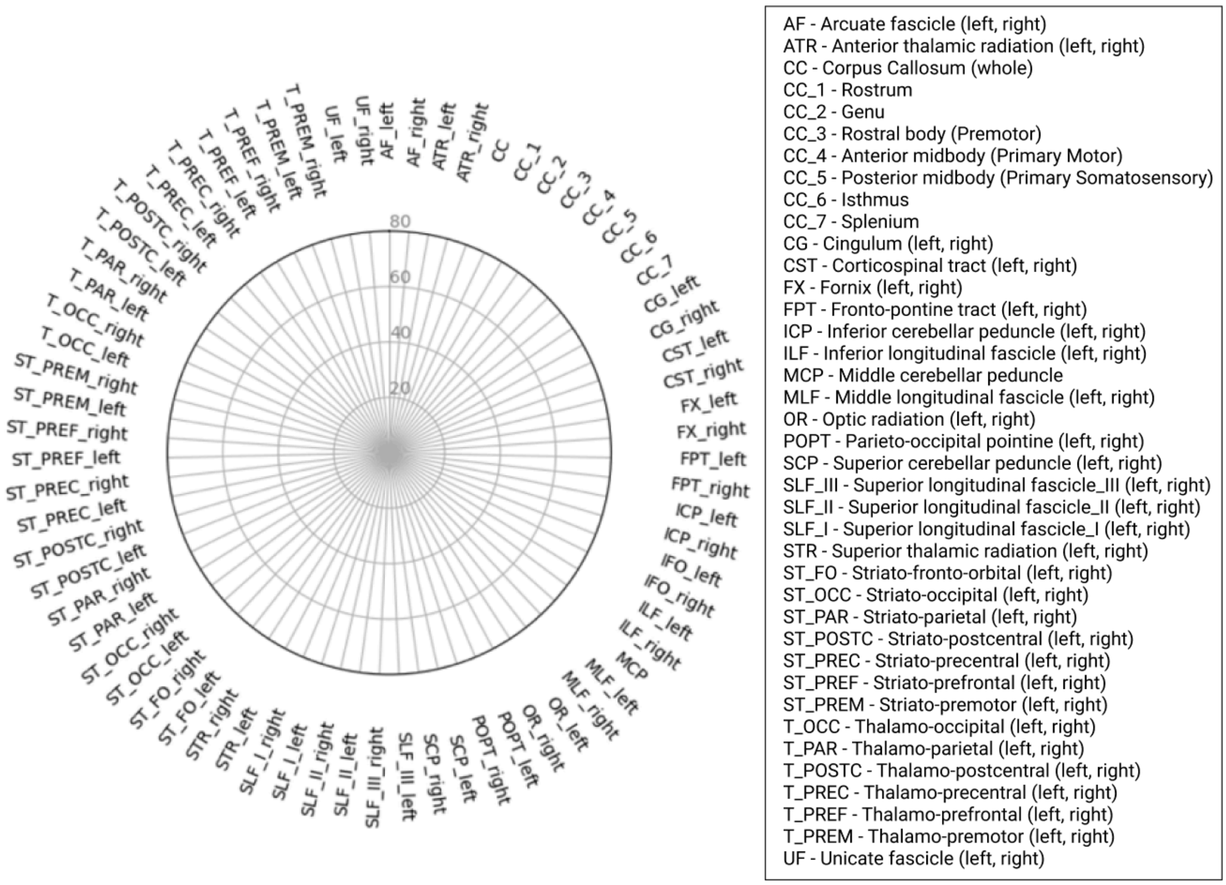


*Supplementary Figure 1.* Example TractLearn radar plots for damaged tract profiles alongside the full list of abbreviations of tracts as described and delineated through TractSeg. Numbers in the plots represent percentage of voxels within tract that is damaged

**References (for supplementary materials only)**


Andersson, J. L., & Sotiropoulos, S. N. (2016). An integrated approach to correction for off-resonance effects and subject movement in diffusion MR imaging. *Neuroimage*, *125*, 1063-1078. doi: 10.1016/j.neuroimage.2015.10.019

Andersson, J. L., Graham, M. S., Zsoldos, E., & Sotiropoulos, S. N. (2016). Incorporating outlier detection and replacement into a non-parametric framework for movement and distortion correction of diffusion MR images. *Neuroimage*, *141,* 556-572. doi: 10.1016/j.neuroimage.2016.06.058

Dhollander, T., & Connelly, A. (2016). A novel iterative approach to reap the benefits of multi-tissue CSD from just single-shell (+b=0) diffusion MRI data. *In: Proceedings of the International Society for Magnetic Resonance in Medicine,* 3010.

Dhollander, T., Clemente, A., Singh, M., Boonstra, F., Civier, O., Dominguez D, J., ... & Caeyenberghs, K. (2021). Fixel-based analysis of diffusion MRI: methods, applications, challenges and opportunities. *Neuroimage, 241,* 118417. doi: 10.1016/j.neuroimage.2021.118417

Dhollander, T., Mito, R., Raffelt, D., & Connelly, A. (2019). Improved white matter response function estimation for 3-tissue constrained spherical deconvolution. *In: Proceedings of the International Society for Magnetic Resonance in Medicine,* 555.

Dhollander, T., Raffelt, D., & Connelly, A. (2016). Unsupervised 3-tissue response function estimation from single-shell or multi-shell diffusion MR data without a co-registered T1 image. *ISMRM Workshop on Breaking the Barriers of Diffusion MRI,* 5.

Egorova, N., Dhollander, T., Khlif, M. S., Khan, W., Werden, E., & Brodtmann, A. (2020). Pervasive white matter fiber degeneration in ischemic stroke. *Stroke*, *51*(5), 1507-1513. doi: 10.1161/STROKEAHA.119.028143

Gajamange, S., Raffelt, D., Dhollander, T., Lui, E., van der Walt, A., Kilpatrick, T., ... & Kolbe, S. (2018). Fibre-specific white matter changes in multiple sclerosis patients with optic neuritis. *NeuroImage: Clinical, 17*, 60-68. doi: 10.1016/j.nicl.2017.09.027

Genc, S., Tax, C. M. W., Raven, E. P., Chamberland, M., Parker, G. D., & Jones, D. K. (2020). Impact of b-value on estimates of apparent fibre density. *Human Brain Mapping, 41*(10), 2583-2595. doi: 10.1002/hbm.24964

Gottlieb, E., Egorova, N., Khlif, M. S., Khan, W., Werden, E., Pase, M. P., ... Brodtmann, A. (2020). Regional neurodegeneration correlates with sleep-wake dysfunction after stroke. *Sleep, 43*(9), zsaa054. doi: 10.1093/sleep/zsaa054

Kellner, E., Dhital, B., Kiselev, V. G., & Reisert, M. (2016). Gibbs-ringing artifact removal based on local subvoxel-shifts. *Magnetic Resonance in Medicine*, *76*(5), 1574-1581. doi: 10.1002/mrm.26054

Raffelt, D. A., Smith, R. E., Ridgway, G. R., Tournier, J. D., Vaughan, D. N., Rose, S., . . . Connelly, A. (2015). Connectivity-based fixel enhancement: Whole-brain statistical analysis of diffusion MRI measures in the presence of crossing fibres. *Neuroimage*, *117*(1), 40-55. doi: 10.1016/j.neuroimage.2015.05.039

Raffelt, D. A., Tournier, J. D., Smith, R. E., Vaughan, D. N., Jackson, G., Ridgway, G. R., Connelly, A. (2017). Investigating white matter fibre density and morphology using fixel-based analysis. *Neuroimage, 144*(A), 58–73. doi: 10.1016/j.neuroimage.2016.09.029

Raffelt, D., Tournier, J. D., Crozier, S., Connelly, A., & Salvado, O. (2012). Reorientation of fibre orientation distributions using apodized point spread functions. *Magnetic Resonance in Medicine*, *67*(3), 844-855. doi: 10.1002/mrm.23058

Raffelt, D., Tournier, J. D., Fripp, J., Crozier, S., Connelly, A., & Salvado, O. (2011). Symmetric diffeomorphic registration of fibre orientation distributions. *Neuroimage*, *56*(3), 1171-1180. doi: 10.1016/j.neuroimage.2011.02.014

Smith, S. M., Jenkinson, M., Woolrich, M. W., Beckmann, C. F., Behrens, T. E., Johansen-Berg, H., ... Niazy, R. K. (2004). Advances in functional and structural MR image analysis and implementation as FSL. *Neuroimage*, *23*, S208-S219. doi: 10.1016/j.neuroimage.2004.07.051

Tournier, J. D., Smith, R., Raffelt, D., Tabbara, R., Dhollander, T., Pietsch, M., ... Connelly, A. (2019). MRtrix3: A fast, flexible and open software framework for medical image processing and visualisation. *Neuroimage, 202,* 116137. doi: 10.1016/j.neuroimage.2019.116137

Wasserthal, J., Neher, P. F., Hirjak, D., & Maier-Hein, K. H. (2019). Combined tract segmentation and orientation mapping for bundle-specific tractography. *Medical Image Analysis*, *58*, 101559. doi: 10.1016/j.media.2019.101559

Wasserthal, J., Neher, P., & Maier-Hein, K. H. (2018). Tractseg-fast and accurate white matter tract segmentation. *Neuroimage*, *183*, 239-253. doi: 10.1016/j.neuroimage.2018.07.070
